# One Health drivers of antibacterial resistance: quantifying the relative impacts of human, animal and environmental use and transmission

**DOI:** 10.1101/2020.06.09.20126433

**Authors:** Ross D. Booton, Aronrag Meeyai, Nour Alhusein, Henry Buller, Edward Feil, Helen Lambert, Skorn Mongkolsuk, Emma Pitchforth, Kristen K. Reyher, Walasinee Sakcamduang, Jutamaad Satayavivad, Andrew C. Singer, Luechai Sringernyuang, Visanu Thamlikitkul, Lucy Vass, OH-DART study group, Matthew B. Avison, Katherine M.E. Turner

## Abstract

**Introduction:** Antimicrobial resistance (AMR), particularly antibacterial resistance (ABR) is a major global health security threat projected to cause over ten million human deaths annually by 2050. There is a disproportionate burden of ABR within lower- and middle-income countries (LMICs), but it is not well understood how ‘One Health’ drivers, where human health is co-dependent on the health of animals and environmental factors, might also impact the burden of ABR in different countries. Thailand’s “National Strategic Plan on Antimicrobial Resistance in Thailand” (NSP-AMR) aims to reduce AMR morbidity by 50% through a reduction of 20% in human antibacterial use and a 30% reduction in animal use starting in 2017. There is a need to understand the implications of such a plan within a One Health perspective that mechanistically links humans, animals and the environment.

**Methods:** A mathematical model of antibacterial use, gut colonisation with extended-spectrum beta-lactamase (ESBL)-producing bacteria and faecal/oral transmission between populations of humans, animals and the environment was calibrated using estimates of the prevalence of ESBL-producing bacteria in Thailand, taken from published studies. This model was used to project the reduction in human ABR (% reduction in colonisation with resistant bacteria) over 20 years (2020-2040) for each potential One Health driver, including each individual transmission rate between humans, animals and the environment, exploring the sensitivity of each parameter calibrated to Thai-specific data. The model of antibacterial use and ABR transmission was used to estimate the long-term impact of the NSP-AMR intervention and quantify the relative impacts of each driver on human ABR.

**Results:** Our model predicts that human use of antibacterials is the most important factor in reducing the colonisation of humans with resistant bacteria (accounting for maximum 72.3 – 99.8% reduction in colonisation over 20 years). The current NSP-AMR is projected to reduce the human burden of ABR by 7.0 – 21.0%. If a more ambitious target of 30% reduction in antibacterial use in humans were set, a greater (9.9 – 27.1%) reduction in colonisation among humans is projected. We project that completely limiting antibacterial use within animals could have a lower impact (maximum 0.8 – 19.0% reductions in the colonisation of humans with resistant bacteria over 20 years), similar to completely stopping animal-to-human transmission (0.5 – 17.2%). Entirely removing environmental contamination of antibacterials was projected to reduce the percentage colonisation of humans with resistant bacteria by 0.1 – 6.2%, which was similar to stopping environment-human transmission (0.1 – 6.1%).

**Discussion:** Our current understanding of the interconnectedness of ABR in a One Health setting is limited and precludes the ability to generate projected outcomes from existing ABR action plans (due to a lack of fit-for-purpose data). Using a theoretical approach, we explored this using the Thai AMR action plan, using the best available parameters to model the estimated impact of reducing antibacterial use and transmission of resistance between populations. Under the assumptions of our model, human use of antibacterials was identified as the main driver of human ABR, with slightly more ambitious reductions in usage (30% versus 20%) predicted to achieve higher impacts within the NSP-AMR programme. Considerable long-term impact may be also achieved through increasing the rate of loss of resistance and limiting One Health transmission events, particularly human-to-human transmission. Our model provides a simple framework to explain the mechanisms underpinning ABR, but further empirical evidence is needed to fully explain the drivers of ABR in LMIC settings. Future interventions targeting the simultaneous reduction of transmission and antibacterial usage would help to control ABR more effectively in Thailand.

## Introduction

Antimicrobials have played an important role in the treatment and prevention of infectious diseases, have enabled food production to intensify and have greatly improved the lives of many millions of people. However, the emergence and spread of antimicrobial resistance (AMR) threatens to undermine this progress, with drug-resistant pathogens projected to cause ten million annual deaths by 2050 [1]. Hence AMR is regarded as a major global health security issue [2], with many member states of the World Health Organization adopting national action plans (with different stages of financing and implementation) in order to tackle the growing threat of AMR [3].

AMR, particularly Antibacterial Resistance (ABR), occurs at the interface of a multifaceted One Health system; human health does not only depend on the human population’s health-related behaviour, but also on industrial, farming and veterinary practices as well as environmental conditions [4]. In the context of ABR, these diverse drivers can be separated into two components: “selection”, predominantly by antibacterial use (ABU) and “transmission” of resistant organisms between each connected compartment on a human-animal-environment axis. In terms of selection, the majority of global ABU is within animals raised for food (73%, [5,6]) and it is generally accepted that ABU in animals drives ABR [7], although the magnitudes of these effects are poorly characterised and are likely to be antibacterial, resistance mechanism and organism specific. However, ABU within human populations (of which up to 50% is unnecessary [8]) is also a fundamental driver of ABR [9]. In terms of transmission, sharing of resistant bacteria between humans, animals and the environment can occur via human-human contact (open community, contact with patients, household transmission, contact in workplaces, travellers), human-animal contact (occupational contact with farm animals, food consumption or preparation, contact with companion animals), animal-animal contact (relating to farming practices, or movement of wild or domesticated animals), and human-environment or animal-environment interfaces (sewage and manure, habitat, drinking water, bathing, leisure activities, food sources and soil) [10]. Crucially, we do not know the full extent to which the listed factors of selection and transmission lead to the currently observed growing prevalence of ABR and increasing incidence of drug-resistant infections.

Selection and transmission of ABR are not entirely independent. ABU drives selection of pre-existing resistant bacteria through population-level mechanisms [11], while simultaneously selecting for successful transmission of resistance into the bacterial population (either via the transmission of mobile genetic elements between microorganisms or through direct transmission of the microorganisms themselves) [7]. Therefore, while ABU in humans and animals, contamination of the environment from those sources or ABU within non-animal agriculture are known to generally increase the prevalence of ABR, the relationship between ABU and resistance is highly complex and dependent on pre-existing bacterial population structures [7]. To account for this, previous studies have suggested that no single *‘silver bullet’* solution exists. Rather, that preventing and reducing the burden of ABR within a One Health system should take a multifactorial, coordinated approach focussing on the specifics of ABU, and the types and prevalence of ABR in each system, while considering the potential interactions within and between compartments [7]. It should also take a multi-sector, transdisciplinary, collaborative approach. One Health is a relatively recent global policy framing of ABR [12] and to date, while animal health has been increasingly included in national policies and action plans, the environment has been given less emphasis [13,14]. One Health approaches are promoted widely in the field, yet the relative contributions of different drivers or the impacts of different interventions are not known, and remain unquantified [10]. Quantification could aid the prioritisation of interventions and refine policy approaches in the inherently complex field of ABR.

While ABR is a global issue [15], there is a disproportionate burden of infectious diseases in lower- and middle-income countries (LMICs) [16]. While these countries experience higher rates of infection (up to three times greater than high income countries [17]), emerging evidence suggests the burden of ABR is greater in LMICs [18] while simultaneously there is limited access to essential antibacterials [8]. Perhaps counterintuitively, research focussed on modelling ABR within lower- and middle-income Southeast Asian countries is vastly underrepresented when compared to European or African studies (eight published models in South-East Asia, 35 in Africa and 42 in Europe [19]), especially when considering that the prevalence of extended-spectrum beta-lactamase (ESBL)-producing *Escherichia coli* (i.e. a key resistance mechanism and pathogen relevant to human and animal health and a sentinel for One Health ABR) in Southeast Asia is high (22% in Southeast Asia compared to 4% in Europe [18]).

Here we focus on the specific national setting of Thailand, an upper-middle income country in Southeast Asia with a high burden of ABR relative to other countries [20] which affects both human health (88 000 infections, 38 000 deaths attributed to ABR per year in 2010) and the economy (direct costs of $70 - 170 million to treat ABR, indirect costs at least $1100 million for morbidity in 2010) [21]. The prevalence of faecal colonisation with ESBL-producing *E. coli* among healthy humans in selected communities in Thailand has grown from 0% in 2004 [22] to 69% in 2010 [23] and 74% in 2012 [24]. In response to the threat posed by rising ABR, the Thai cabinet implemented their first five-year policy in 2016, the “National Strategic Plan on Antimicrobial Resistance in Thailand” (NSP-AMR) running until 2021 [25]. The NSP-AMR reflects the strategic objectives of the WHO Global Action Plan [3] and aims to reduce AMR morbidity in Thailand by 50% through 20% reductions in AMU in humans, 30% reductions in AMU in animals and 20% increases in public knowledge on AMR (including awareness of AMU) by 2021 [25].

We aim to quantify the relative contributions to the human ABR burden (% colonisation with resistant bacteria) of human, animal and environmental factors (including ABU and transmission of ABR bacteria) within a One Health system in Thailand. We propose a simple mathematical framework for the spread of ABR between these compartments from which we will explore and assess national interventions aimed at reducing ABR. Our objective is to assess and compare a wide variety of One Health drivers and provide insights into the multifaceted problem of ABR. This simple model is intended to stimulate further discussion on how best to reduce the burden of ABR in human populations and to provide the much needed first step in providing a workable One Health modelling framework.

## Methods

We built a compartmental model of ordinary differential equations (ODEs) to describe the relationship between resistant bacteria in three compartments: humans, animals and the environment. We considered the fraction of all humans colonised with resistant bacteria (H), the fraction of animals with resistant bacteria (A) and the fraction of environmental samples with resistant bacteria (E) based on the framework of a previously published model of animal-human transmission [26]. Here, we assume that the bacteria are ESBL-producing *E. coli*, and human/animal colonisation is assumed to be of the gut and via the faecal-oral route. Given our use of a sentinel pre-evolved resistance mechanism, *de novo* selection of resistance within a compartment is not considered significant.

We assumed that resistance develops from two sources; antibacterial use (*Box 1A*, which selects resistant bacteria already present in the host) and transmission from other compartments (*Box 1B*, which is also dependent on antibacterial use in those compartments).

First, resistance develops in humans from exposure to antibacterials, proportional to their usage in medicine Λ_H_, the rate at which humans are colonised with resistant bacteria, *γ*, and the fraction of humans not already colonised (1−*H*). In a similar fashion, resistance develops in animals proportional to antibacterial usage in companion animal, veterinary and farm practices Λ_A_, the rate at which animals are colonised with resistant bacteria, *γ*, and the proportion of animals not already colonised (1− *A*)(*Box 1A*).

Λ_E_represents the presence of antibacterial in the environment (derived from factories, pollution, wastewater etc), while (1− *E*)represents the proportion of environmental samples negative for ABR bacteria. We assumed that the prevalence (percent of humans, animals or environmental samples that have resistant bacteria) of ABR bacteria declined at rates *μ*_*H*_, *μ*_*A*_ and *μ*_*E*_ within humans, animals and the environment respectively.

**Box 1:**
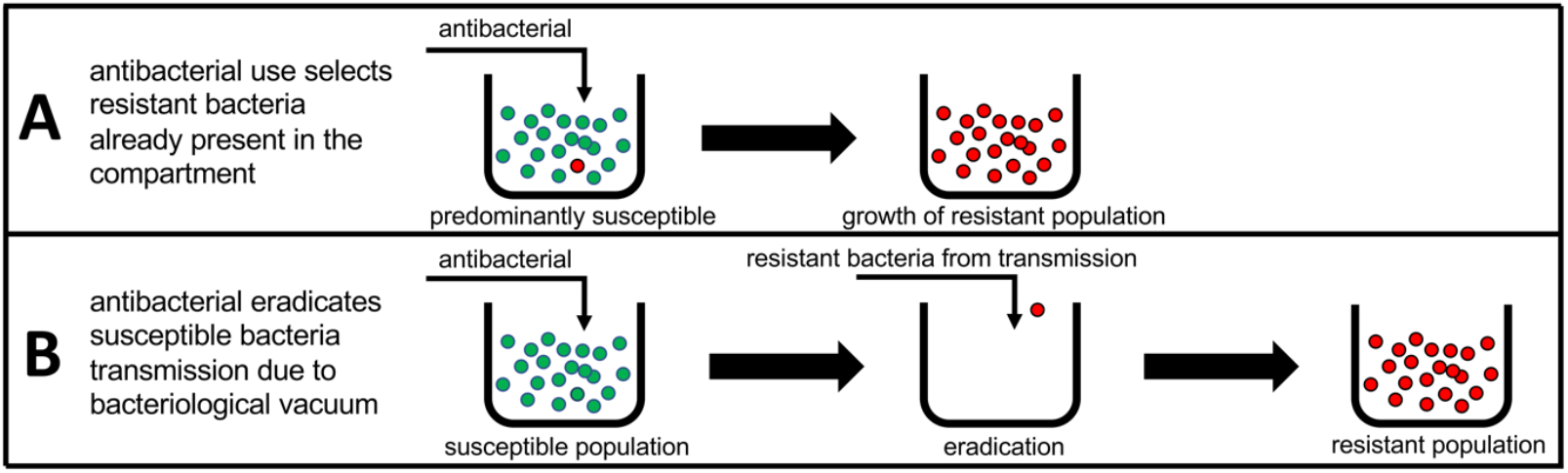
Two mechanisms of resistance driven by antibacterial exposure. First, antibacterial use selects resistant bacteria already present in the compartment, and second that antibacterial use can remove susceptible populations making transmission more likely (it is more likely for colonisation with resistant bacteria to occur when the native flora have been reduced/removed).

Second, we assumed that humans negative for ABR bacteria are exposed to and acquire resistant bacteria from other humans at rate *β*_*HH*_ (representing transmission in the open community and from patients, farm workers and other high-risk groups). Acquisition in this context means the combined effects of stable colonisation with a resistant bacterium following transfer, and infiltration of mobile resistance genes (e.g. via plasmid) into the existing bacterial flora following transient colonisation with a transferred resistant bacterium. Similarly, animal-to-animal acquisition of resistance occurs at rate *β*_*AA*_, and animal-to-human transfer of resistance occurs at rate *β*_*AH*_ (representing contact with farm animals, contact with companion animals and food consumption). Human-to-animal transmission of resistance occurs at rate *β*_*HA*_, however this rate is smaller than the animal-to-human rate *β*_*AH*_ (as livestock to human jumps occur more frequently over evolutionary history than *vice versa* [10]). Similarly, the rates of acquisition of resistance occur at *β*_*EH*_ for environment-to-human (drinking water, non-animal food sources, swimming and bathing in freshwater), *β*_*EA*_ for environment-to-animal (habitat, food sources, drinking water), *β*_*HE*_ for human-to-environment (transfer through sewage) and *β*_*AE*_ for animal-to-environment (manure, or composting of dead animals). We assumed that transmission from the environment to animals was greater than that of the environment to humans (*β*_*EA*_>*β*_*EH*_), as a previous study showed that there were higher proportions of shared bacterial genera in wastewater and animals, as opposed to wastewater and humans [27]. We also assumed that the transmission within populations of humans and animals is greater than the transmission between these populations and the transmission from the environment to these populations (*β*_*HH*_>*β*_*AH*_,*β*_*EH*_ and *β*_*AA*_>*β*_*HA*_,*β*_*EA*_).

We assumed that all rates of transmission are proportional to exposure with antibiotics, due to treatment eradicating susceptible bacteria in the host thereby enabling colonisation by incoming resistant bacteria (*Box 1B*, [11]). Therefore, the dynamics for the fraction of humans, animals and the environment with resistant bacteria is represented by the following ordinary differential equation model (Figure 1):

**Figure 1:**
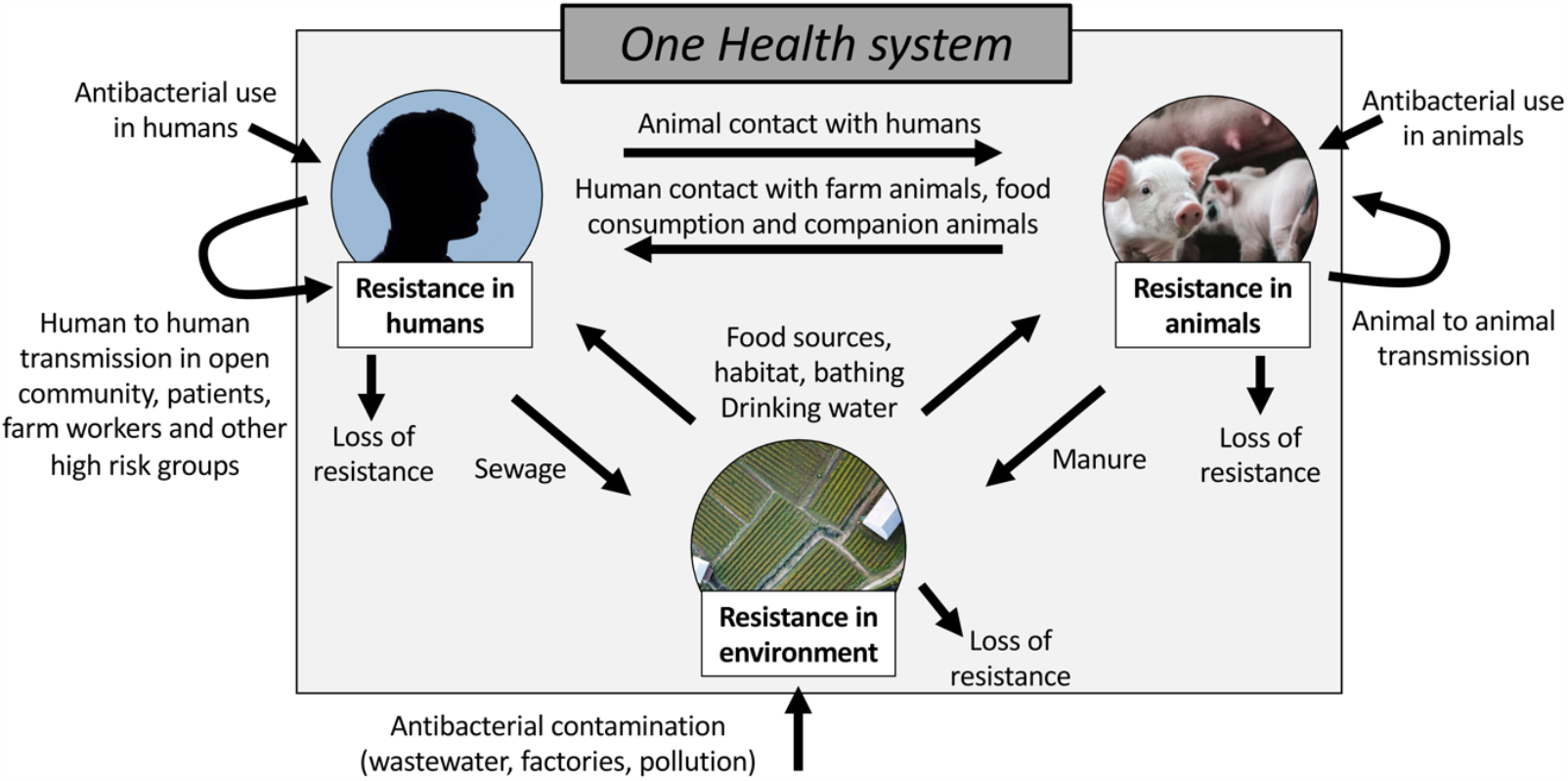
Model schematic for human-animal-environment transmission of ABR in the One Health setting.

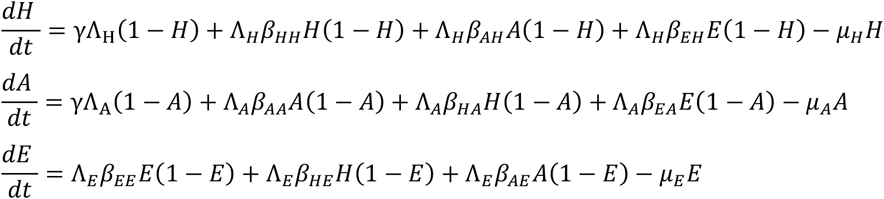

For example, the rate of change over time for the fraction of humans with resistant bacteria 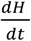= the rate at which susceptible humans become colonised with resistant bacteria *γ*Λ_H_(1−*H*)+ the transmission from humans with ABR to susceptible humans Λ_*H*_*β*_*HH*_*H*(1−*H*)+ the transmission from animals with ABR to susceptible humans Λ_*H*_*β*_*AH*_*A*(1−*H*)+ the transmission of resistant bacteria in the environment to susceptible humans Λ_*H*_*β*_*EH*_*E*(1−*H*)– the loss of resistance *μ*_*H*_*H* at each time point.

We assumed that Λ_*A*_>Λ_*H*_ as the majority of global ABU is within animals raised for food (73%, [5,6]). We further assumed that ABU is greater in humans when compared to the amounts present in the environment Λ_H_>Λ_*E*_, and that the rates of loss of resistance are equal in all three settings (*μ*_*H*_=*μ*_*A*_=*μ*_*E*_) in the absence of data on decay of resistance within different compartments.

The model was coded and numerically simulated using R v.1.2.5019. ODEs were solved using functions *deSolve* and *ode*.

We identified estimates (Table 1) for the prevalence of gut colonisation with ESBL-producing *E. coli* in Thailand for healthy humans across an eight-year timeframe (2004 – 2012) [22–24,28,29], estimates of sample level positivity for resistant bacteria in rectal swabs from animals and fresh food from Thailand in 2012-2013 [24], and environmental estimates of the proportion of bacteria that are resistant in Thailand from canal water sources [24], stagnant water on food animal farms, and liquid from hospital wastewater treatment tanks [30]. These data were used to calculate lower and upper bounds (95% credible intervals) in order to calibrate our model to a national-level Thailand-specific setting.

**Table 1:** Parameters and fitting metrics used in the model (summary of 95% confidence interval uncertainty ranges). HH = human-human, AH = animal-human, AA = animal-animal.

| Model Parameters | Parameter value (95% confidence interval) | Source |
| --- | --- | --- |
| Antibacterial usage |  |  |
| Animals | 0 – 100% | Majority of antimicrobials used in animals raised for food (73%, [5,6]) |
| Human | 0 – 100% of animal use |  |
| Environment | 0 – 100% of human use |  |
| Transmission of ABR |  |  |
| Humans to humans | 0.0 - 100.0 % | Explore full parameter space |
| Animals to humans | 0.0 – 100.0% of HH | Fraction of HH transmission |
| Environment to humans | 0.0 – 100.0% of HH | Fraction of HH transmission |
| Animals to animals | 0.0 – 100.0% | Explore full parameter space |
| Humans to animals | 0.0 – 100% of AH and AA | Assume wide range, but less than AH [10] and AA transmission |
| Environment to animals (habitat/ drinking water) | 0.0 – 100.0% of AA | Fraction of animal-animal transmission |
| Humans to environment (sewage) | 0.0 – 100.0% | Explore full parameter space |
| Animals to environment (manure) | 0.0 – 100.0% | Explore full parameter space |
| Carriage of resistance |  |  |
| Rate of loss of resistant bacteria after 12 months | 0-100% | Explore full parameter space |
| Fitting Parameters | Value (95% confidence interval) | Source |
| Prevalence of colonisation with ESBL-producing organisms |  |  |
| Humans |  |  |
| 2004 | 0.0 – 0.0% | 0/120 healthy adults in Thailand [22] |
| 2008 | 46.7 – 62.1% | 87/160 healthy asymptomatic volunteers in Thailand [28] |
| 2009 | 35.2 – 44.3% | 177/445 healthy asymptomatic volunteers in Thailand [29] |
| 2010 | 64.9 – 73.7% | 289/417 door to door sampling among healthy individuals in Thailand [23] |
| 2012 | 71.4 – 78.5% | 430/574 healthy workers in a food factory and food animal farm in Thailand [24] |
| Animals | 11.1 – 60.3% | Upper bound from rectal swabs in pigs (241/400), lower bound from fresh food (12/54) in Bangkok and East/North Thailand [24] |
| Rectal swabs of animals, 2012-2013 |  |  |
| Pig | 55.4 – 65.0% | 241/400 in East/North Thailand [24] |
| Broiler | 26.9 – 48.1% | 30/80 in East/North Thailand [24] |
| Laying Hen | 0.0 – 7.7% | 2/61 in East/North Thailand [24] |
| Fresh pork meat | 0.0 – 25.6% | 2/18 in East/North Thailand [24] |
| Fresh food, 2012-2013 |  |  |
| Chicken | 4.9 – 52.2% | 4/14 in Bangkok & central [24] |
| Pork | 28.1 – 78.6% | 8/15 in Bangkok & central [24] |
| Beef | 0.0 – 0.0% | 0/11 in Bangkok & central [24] |
| Fish | 0.0 – 0.0% | 0/14 in Bangkok & central [24] |
| Environment | 0.0 – 24.7% | Lower bound from water in Bangkok and central province [24] and upper bound from stagnant water on food animal farms [24] |
| Canal water, 2012-2013 | 0.0 – 19.3% | 1/15 in Bangkok and central [24] |
| Stagnant water on food animal farms, 2012-2013 | 0.0 – 24.7% | 3/25 in Bangkok and central [24] |
| Effluent fluid after treatment with chlorine prior to draining into a public water source from hospitals | 2.4 – 17.6% | 6/60 in public hospitals in Thailand [30] |

We assumed the widest possible range for parameters with no prior data and extrapolated based on the hierarchy of transmission (e.g. human-human transmission is greater or equal to animal-human transmission). Therefore, we allowed human-human transmission to take any possible value (from 0 – 100%), while animal-to-human transmission is a fraction of this value (0 – 100% of human-human). We explored the full range of transmission for animal-animal, animal-environment and human-environment, while environment-human is a fraction of human-human, and human-animal and environment-animal are a fraction of the animal-animal rate (Table This method ensures that we explored every possible scenario while maintaining the structure of the model and the hierarchy of the separate transmission rates.

Similarly, we allowed ABU within animals to vary between the minimum and maximum values (0 – 100%), while human use is a fraction of this (less than or equal to animal use), and environmental use or contamination is a fraction of human use (Table 1).

## Results

From 1 000 000 simulated parameter sets, 241 agreed well with the lower and upper bounds (Table 1) for the percentage of humans, animals and environmental samples colonised with resistant bacteria in Thailand (Figure 2). The fitting bounds for the percentage of humans with resistant bacteria was underestimated in 2009, perhaps due to discrepancy in sampling methodology or a different sampling cohort [29]. The full description of the prior and posterior parameter ranges can be found in the supplementary information (Figure S1).

**Figure 2:**
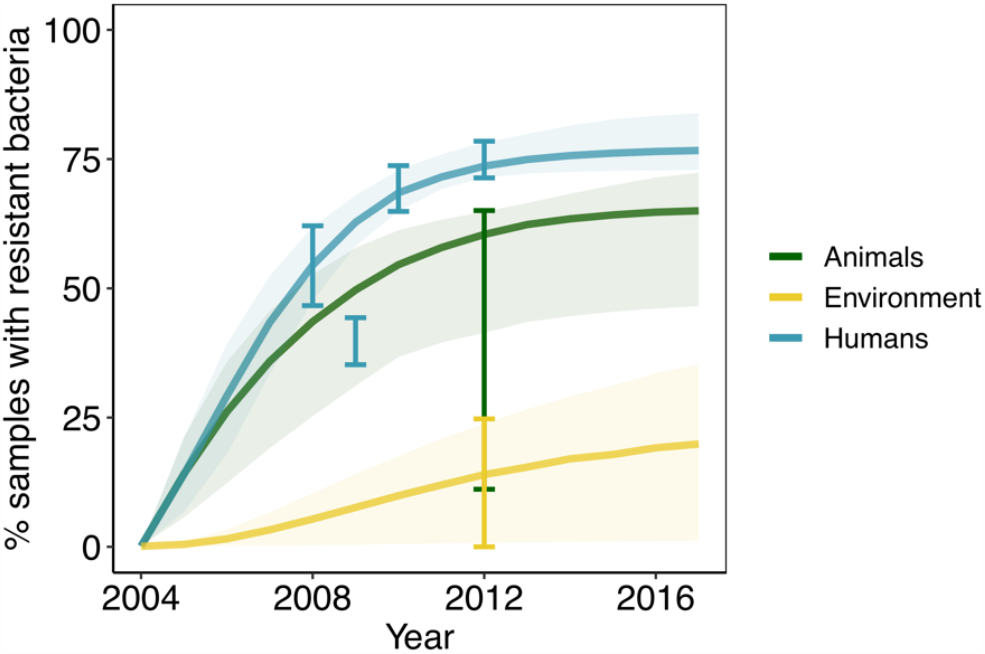
Comparison of the model projections for animals, humans and the environment for the best 241 model fits (from 1 million samples). Error bars indicate data from which the model was calibrated (3/4 for humans, aside from 2009) and shaded areas indicate the 95% confidence interval among all fits. Model parameters and their uncertainty ranges can be found in Table 1.

### The National Strategic Plan on AMR in Thailand

Table 2 shows the potential predicted impact of reduction in colonisation with resistant bacteria in humans via interventions starting in 2020 and running for 20 years until 2040 (compared to the current standard of care). Over 20 years (2020-2040), the NSP-AMR in Thailand (which aims to reduce antimicrobial use in humans by 20%, animals by 30% and increase public knowledge of AMR equating to an increase in all sanitary practices e.g. handwashing, safe drinking water, sewage disposal, waste water treatment plants by 20%) was estimated to reduce the number of humans colonised with resistant bacteria by 12.7% (95% credible interval: 7.0 – 21.0%, Table 2) from 2020 values by 2040. The NSP-AMR supplemented with an additional reduction in human-based ABU (to 30%) would reduce the burden of resistance (% colonised with resistant bacteria) within humans to a greater extent: to 17.1% (9.9 – 27.1%, Table 2) of 2020 values. The NSP-AMR supplemented by an additional increase in sanitary practices (to 30%, we predict an impact similar to reducing ABU by 30%: a reduction of 16% (8.6 – 27.4%, Table 2) from 2020 values.

**Table 2:** Reduction in colonisation with resistant bacteria in humans between 2020 - 2040 for different potential interventions. The National Strategic Plan on Antimicrobial Resistance (NSP-AMR; 2017-2021) in Thailand aims to reduce antibiotic use (ABU) in humans by 20%, animals by 30% and increase sanitary knowledge by 20% (in 2021). We investigate other hypothetical interventions aiming to reduce ABU in humans and animals and other transmission related interventions.

| *Intervention or scenario* | *Median reduction in colonisation with resistant bacteria in humans (95% credible interval) 2020-2040* |
| --- | --- |
| NSP-AMR, Thailand | 12.7 (7.0 – 21.0) |
| NSP-AMR with 30% reduction in human ABU | 17.1 (9.9 – 27.1) |
| NSP-AMR with 30% increase in sanitary practices | 16.0 (8.6 – 27.4) |
| 50% reduction in all transmission only | 15.0 (6.9 – 29.8) |
| 50% reduction in water related transmission only | 1.3 (0.0 – 3.6) |
| 20% reduction in human ABU with 50% reduction in human-human transmission | 14.2 (7.7 – 22.7) |
| 20% reduction in human ABU with 95% reduction in transmission from sewage/manure | 7.8 (4.8 – 12.4) |

Without any ABU reduction, reducing all transmission rates in our model to 50% of their original value was predicted to reduce the number of humans carrying resistant bacteria to 15% (6.9 – 29.8%, Table 2) by 2040. However, the impact of reducing only the transmission rates relating to water and the environment (human and animal transmission to and from the environment) on colonisation in humans was relatively low (1.3% reduction, 0.0 – 3.6%, Table 2). A 20% reduction in human ABU together with a 50% reduction in human-human transmission were projected to reduce the human burden of resistance by 14.2% (7.7 – 22.7%, Table 2), while a 20% reduction in human ABU with a 95% reduction in sewage or manure transmission was projected to reduce the burden of human resistance by 7.8% over 20 years (4.8 – 12.4%, Table 2).

Figure S2 shows the potential impact of simultaneously reducing ABU in humans and animals (in intervals of 10%; 0, 10, 20, 30, 40 and 50%) and increasing sanitary practices (transmission remains at current levels, reducing transmission by 25%, 50% and 75%). In the absence of any sanitary interventions aiming to reduce transmission, achieving the NSP-AMR targets for ABU reduction would result in an impact of 7.3% reductions in human colonisation of resistant bacteria by 2040. Increasing sanitary practices to 25% would result in a 2.1-fold reduction in resistant bacteria colonisation in humans (to 15.2%), while increasing sanitary practices by 50% and 75% would result in larger 3.5-fold (to 25.6%) and 7.2-fold (to 52.3%) reductions in the presence of the current Thailand NSP-AMR targets.

### Which factors contribute the most to human ABR

The maximum potential reduction in colonisation with resistant bacteria for humans from single parameters in the model are summarised in Table 3 (more detail available in Table S1). While removing human ABU (0% ABU in humans) has the highest potential impact (72.3 – 99.8% reduction in colonisation of resistant bacteria over 20 years), reducing human-to-human transmission (7.4 – 38.8%), animal ABU (0.8 – 19.0%) and animal-to-human transmission (0.5 – 17.2%) all have considerable potential impact. Environmental contamination and environment-to-human transmission were both predicted to have a smaller impact (<6.2%).

**Table 3:** The maximum reduction in resistant bacteria within human populations when accounting for the effects of usage and transmission parameters. The maximum reduction can be achieved by totally stopping human antibacterial use and human-human transmission, but animal and environmental factors can still contribute significantly to reducing the burden of ABR within humans.

| *Parameter* | *Median reduction in colonisation with resistant bacteria in humans (95% credible interval) 2020-2040* |
| --- | --- |
| Human ABU | 72.3 – 99.8% |
| Human-to-human transmission | 7.4 – 38.8% |
| Animal ABU | 0.8 – 19.0% |
| Animal-to-human transmission | 0.5 – 17.2% |
| Environment contamination of AB | 0.1 – 6.2% |
| Environment-to-human transmission | 0.1 – 6.1% |

Table S1 shows the maximum potential reduction in colonisation with resistant bacteria in humans for 30 scenarios, ranked by their relative contributions to human ABR. The top four ranked scenarios were all related to human ABU (96% reduction in colonisation over 20 years in humans). The next highest factors were the rate of loss of resistant bacteria in humans (64.9%), followed by no transmission events (43.7%), simultaneous human-human/human-environment/human-animal transmission (19.5%) and human-human transmission alone (17.3%).

The relative attributable impact on the burden of colonisation of resistant bacteria within human populations of human ABU compared to use in animals was determined to be 16:1 (16 times more impact could be achieved through reducing ABU in humans rather than animals, Table S1). This ratio decreased to 13:1 when accounting for all animal-based transmission. Likewise, the relative impact on human ABR of environmental contamination of antibacterials was a ratio of 55:1 (50:1 when accounting for environmental transmission events) but decreased to 10:1 when combined with animal ABU and all environmental and animal-based transmission. When comparing the relative impact of the human AMR burden to human-human transmission, the ratio was 6:1 (six times more impact of reducing human ABU compared to reducing human-human transmission), and comparing to all transmission routes in the model resulted in a ratio of 2:1 (two times more impact of reducing human ABU compared to reducing all transmission).

### Which factors contribute the most to animal and environmental ABR

The top ranked scenarios for both animals and the environment were all related to ABU in animals and antibacterial contamination of the environment, respectively (Table S1). Interestingly, removing all human usage (human ABU = 0) would result in a 3.8% reduction in animals colonised with resistant bacteria, but a much greater 34.6% of the resistant contamination of bacteria in the environment. Removing animal ABU (animal ABU = 0) resulted in a 96.4% reduction in animals and 30.8% reduction in the environment, while removing environmental antibacterial contamination resulted in 1.4% and 96.8% reductions in colonisation in animals and the environment, respectively (Table S1).

## Discussion

Prior to this study, an AMR literature review found that only 2% of published models (*five from a total of 273*) considered human and animal transmission concurrently, and no published model considered a third environmental setting [19]. To our knowledge, therefore, our study is the first to consider the One Health human-animal-environment axes of ABR. This is an especially important factor to consider as human ABU, animal ABU and environmental antibiotic contamination have all been shown to increase the prevalence of ABR [6,10,31,32]. We propose this simple model as a first step in understanding the complex picture [7] of One Health ABR, but our model framework includes assumptions which should be recognised, and thus these results should be interpreted in light of these assumptions.

First, prevalence data we used in the fitting of our model came from the six separate studies available in the literature – each with varying methodologies and cohorts. We did, however, explore the reliability of each of these national Thai estimates from 2004 to 2013 with a 95% confidence interval calculated for each reported data point – ensuring that we accounted for differences between these studies. There is a paucity of data on the transmission of bacteria between the different compartments in any One Health system, and no information at all for the situation in Thailand. Instead we assumed a hierarchy of transmission based on other published studies from other countries. For example, we used a study of shared bacterial genera in Beijing, China to inform the assumption that environment-to-animal transmission is greater or equal to that of environment-to-human transmission. We also used a wide range of potential estimates (with 95% confidence intervals) and explored every possibility of all underlying parameters.

Our model primarily considers ABR bacteria transmitted via the faecal-oral route and those carrying mobile resistance genes (i.e. ESBL-producing *E. coli*, which is a very common sentinel for ABR in a One-Health context) [24]. Whilst bacteria transmitted via faecal oral route make up a high proportion of the World Health Organisation list of priority ABR pathogens [33], other types of resistant bacteria (e.g. MRSA) do not have an environmental reservoir, and indeed require close physical contact in order to transmit – so this model does not capture those resistance mechanisms of *all* resistant bacteria. Instead we focus on those bacteria which simultaneously affect all aspects of the One Health network [24]. In addition, the lack of fit-for-purpose data to inform this model may have skewed our results to favour humans as the most important factor (as this is where the data is most rich), and future data may clarify the complete role of animals and the environment in human ABR. Future models should extend our framework to these aspects and especially consider the setting-specific features of the population and their behaviour. This would result in a better understanding of the One Health drivers of ABR in human populations.

Widespread concern around the contribution of animal antibacterials to human resistance is growing, perhaps due to the majority of use, globally, occurring within this sector [5,6], but the benefits of curtailing their use on human health remain unquantified [10]. Here, the potential absolute impact of limiting the use of antibacterials in animals was predicted to be far less than then limiting human medical use (16 times more impact achieved through reducing ABU in humans when compared to animals) but this ratio was decreased when transmission between animals and humans was accounted for (13 times more impact), and further decreased when accounting for environmental contamination (10 times more impact). This result suggests that while animal ABU has been highlighted as a major driver within human ABR [5], it is far more effective to reduce human ABU in the first instance. However, reducing human ABU may not be feasible in many cases – and indeed removing animal use entirely would also contribute to minimising the development of ABR within animals (72 – 100%, Table S1) with substantial knock-on implications for human health (up to an impact of 20% reductions by 2040). This suggests that animal antibacterials are still an important driver for human resistance (albeit not as important as human use). Removing animal use entirely is also not realistic if animals are being used to produce food, and these results should be considered carefully for welfare reasons, even when making minor reductions in use (this model does not suggest we should reduce anything by 100%, rather that these results indicate which areas of the One Health system yield maximum impact).

Limiting human ABU was six times as effective as reducing human-human transmission and twice as effective as reducing all transmission events. This suggests that interventions targeting reductions in ABR within human populations should also focus on improved hygiene and infection control (particularly for humans) in addition to curtailing ABU.

Overall, therefore, we predict that the most effective method of reducing the burden of resistant bacteria in humans is to combine reductions in ABU while simultaneously reducing transmission events between humans, animals and the environment, reinforcing the need for One Health approaches that consider all three. This finding agrees with the results of other studies: one such study found that animal ABU alone had little impact on levels of human ABR [26] while another found that resistance in hospitals could be better prevented by interventions simultaneously targeting transmission and antibacterial exposure [32] - however none of these studies considered ABU within a human-animal-environment One Health framework.

The current NSP-AMR in Thailand (2017-2021) aims to reduce ABU and transmission simultaneously, which according to our results is the most efficient way of reducing the burden of ABR in humans. We show that successfully achieving and maintaining current targets [25] until 2040 would result in a reduction of 7.0 – 21.0% in the number of people carrying resistant bacteria (assuming a 20% increase in sanitary practices to decrease transmission through ABR public knowledge). This impact is reasonable but could be improved by further reductions of human ABU (from 20% to 30%) (NSP-AMR; 9.9 – 27.1%). Alternatively, halving all transmission events alone (which depend on sanitary practices) without NSP-AMR targets being met was almost as effective in reducing ABR (6.9 – 29.8%). This shows that there are multiple alternatives which could strengthen the current NSP-AMR in Thailand. One such alternative is a combined approach: the current NSP-AMR with 50% reductions in transmission (compared to no change in sanitary practices) would be 3.5-times as effective in reducing human colonisation with resistant bacteria (median 25.6% reduction over 20 years). This shows that the general impact of restricting ABU can be greatly enhanced in the context of reduced transmission across various settings.

While our model is initially developed for the context of Thailand (the case study for model parameterisation), these results are potentially generalisable to any country or region with a high prevalence of resistant bacteria, including other Southeast Asian countries (22% prevalence of faecal colonisation with ESBL producing *Enterobacteriaceae*), African countries (22%), West Pacific countries (46%) or eastern Mediterranean countries (15%) [18]. However, it would be unreasonable to assume that each of these geographical regions share the exact parameters used in our Thailand study. For example, almost half of the Thai population are employed in the agricultural sector [34] and 30.1% of people use water from natural sources [35]; while some countries have similar demography, it will be important to collect parameter estimates specific for each country and region. However, ABR research in LMICs is characterised by data gaps as well as variability in data reliability, sharing and capacity [36]. In the absence of such data [36], this initial model may be used as a first step in understanding and evaluating other LMICs’ ABR strategies. Ideally, future studies would obtain country-specific data on ABR (particularly for LMICs) or regional data from these countries. Then, a similar data-driven approach could be used to predict and forecast future ABR interventions with higher degrees of certainty in each specific region.

Our model makes some important predictions which have direct implications for human health in the context of ABR. Our conceptual model identified that human antibacterials are the primary driver in human ABR, but that there are many such interacting drivers which, if targeted by the correct interventions, could have large implications for the wider ABR problem. Interventions which focus on reducing ABU in humans can yield much greater impact when run in parallel to improved hygiene and sanitation interventions. Future work is needed to develop this model framework and to capture high-resolution data on transmission events between humans, animals and the environment, and to quantify the effects of ABU within animals and the environment on human health. This model has allowed estimation of the impact of the Thai NSP-AMR and has suggested where greater ambition in its targets could significantly increase its potential impact on ABR. We anticipate that the results of this modelling study will stimulate further discussion on One Health interventions within Thailand and across other LMICs.

## Data Availability

All data is from publicly available sources and are listed in Table 1

## Acknowledgements

This work was funded by grant MR/S004769/1 to M.B.A. from the Antimicrobial Resistance Cross Council Initiative supported by the seven United Kingdom research councils and the National Institute for Health Research.

## Supplementary Information

**Figure S1:**
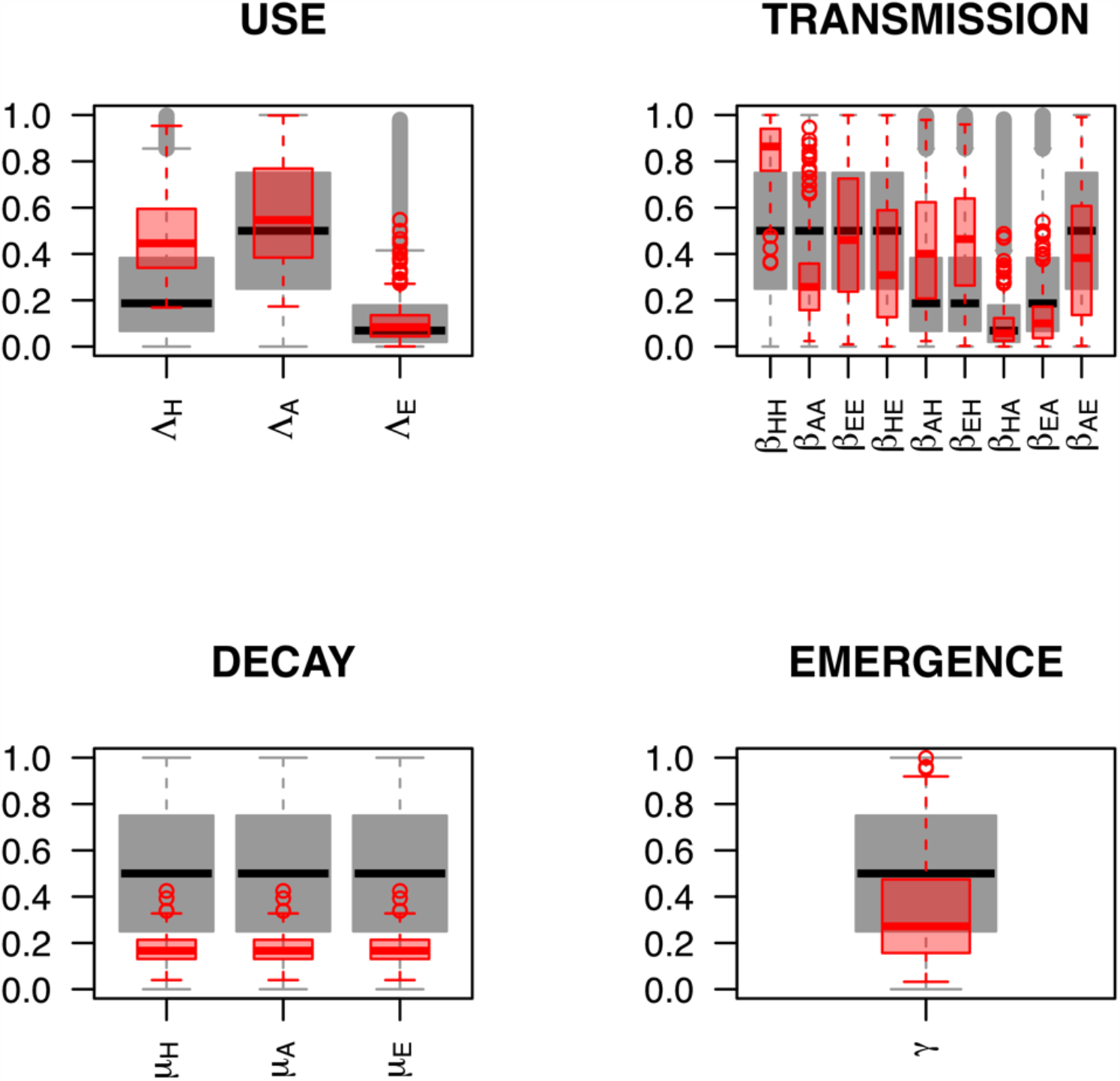
Parameter prior and posterior distributions. Grey boxes represent the search range/prior distribution of parameters (taken from Table 1), while red boxes represent the posteriors from the best 241 hundred fits (from a sample of 1 million), with interquartile range shaded, median, and minimum and maximum values.

**Figure S2:**
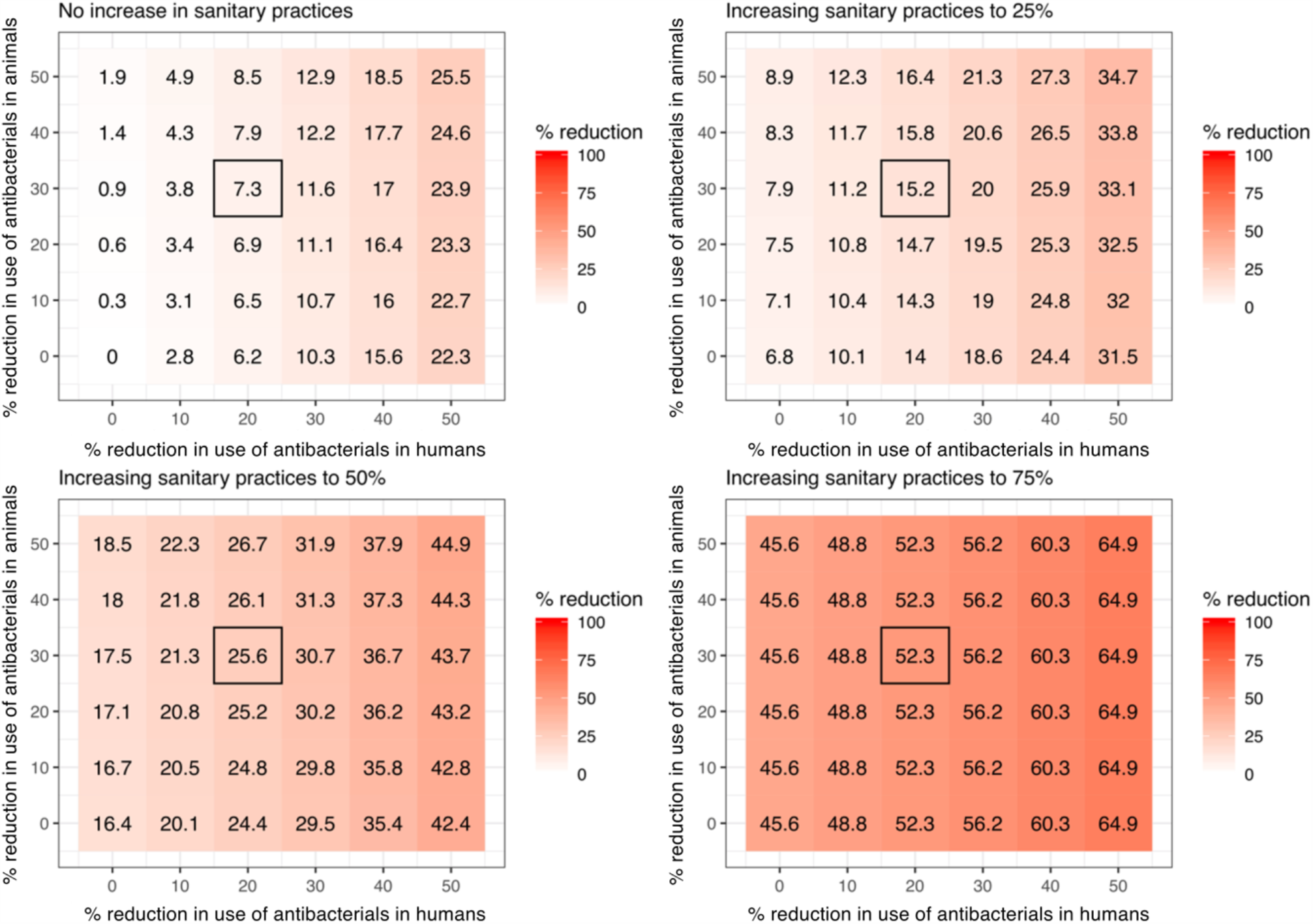
The relationship between reducing transmission via increasing knowledge of sanitary measures and antibacterial use in humans and animals. Black square indicates the current National Strategic Plan on Antimicrobial Resistance (2017-2021) (NSP-AMR) targets for 0, 25, 50, 75 and 100% reductions in sanitary related transmission. With 20% reductions in use in humans, and 30% in animals, alongside no reduction in transmission related to education or prevention would reduce ABR in humans by 7.3%. A 25% increase in sanitary practices would result in 2.1-fold higher reductions in human ABR, 50% would result in 3.5-fold higher impact (25.6% reduction in human ABR) and 75% increases in sanitary practices would result in 7.2-fold higher impact (52.3% reduction in human resistance).

**Table S1:** Reduction in colonisation with resistant bacteria in humans, animals and the environment from 2020-2040, in order of impact on human burden of resistant bacteria for combinations of use and transmission and loss of resistance.

| *Intervention* | *Parameters* | *Median reduction in colonisation with resistant bacteria in humans (95% credible interval) 2020-2040* | *Median reduction in colonisation with resistant bacteria in animals (95% credible interval) 2020-2040* | *Median reduction in colonisation with resistant bacteria in the environment (95% credible interval) 2020-2040* |
| --- | --- | --- | --- | --- |
| No antibacterial use and no transmission | $\Lambda_H = \Lambda_A$<br>$= \Lambda_E = 0$<br>$\beta = 0$ | 96.4 (72.3 – 99.8%) | 96.4 (72.3 – 99.8%) | 96.8 (82.3 – 99.8%) |
| No antibacterial use | $\Lambda_H = \Lambda_A$<br>$= \Lambda_E = 0$ | 96.4 (72.3 – 99.8%) | 96.4 (72.3 – 99.8%) | 96.8 (82.3 – 99.8%) |
| No human antibacterial use | $\Lambda_H = 0$ | 96.4 (72.3 – 99.8%) | 3.8 (0.4 – 23.5%) | 34.6 (3.2 – 82.0%) |
| No human antibacterial use and no human to human, human to environment, human to animal transmission | $\Lambda_H = 0$<br>$\beta_{HH} = \beta_{HE}$<br>$= \beta_{HA} = 0$ | 96.4 (72.3 – 99.8%) | 4.5 (0.6 – 25.8%) | 42.4 (4.2 – 91.1%) |
| Increasing rate of loss of resistance in humans to max | $\mu_H = 1$ | 64.9 (45.8 – 78.0%) | 2.7 (0.4 – 11.0%) | 25.0 (2.7 – 56.0%) |
| No transmission | $\beta = 0$ | 43.7 (17.2 – 81.4%) | 23.9 (3.2 – 74.4%) | 96.8 (82.3 – 99.8%) |
| No human to human, human to environment, human to animal transmission | $\beta_{HH} = \beta_{HE}$<br>$= \beta_{HA} = 0$ | 19.5 (8.3 – 41.7%) | 4.5 (0.6 – 25.8%) | 42.4 (4.2 – 91.1%) |
| No human to human transmission | $\beta_{HH} = 0$ | 17.3 (7.4 – 38.8%) | 0.7 (0.1 – 4.3%) | 6.0 (0.6 – 23.5%) |
| No antibacterial use in animals and environment, no animal to animal, animal to human, animal to environment, no environment to animal, environment to human, environment to environment transmission | $\Lambda_A = \Lambda_E = 0$<br>$\beta_{AA} = \beta_{AH}$<br>$= \beta_{AE} = 0$<br>$\beta_{EA} = \beta_{EH}$<br>$= \beta_{EE} = 0$ | 9.2 (1.7 – 24.1%) | 96.4 (72.3 – 99.8%) | 96.8 (82.3 – 99.8%) |
| No antibacterial use in animals and environment, no animal to animal, animal to environment, no environment to animal, environment to environment transmission | $\Lambda_A = \Lambda_E = 0$<br>$\beta_{AA} = \beta_{AE} = 0$<br>$\beta_{EA} = \beta_{EE} = 0$ | 8.3 (1.4 – 23.2%) | 96.4 (72.3 – 99.8%) | 96.8 (82.3 – 99.8%) |
| No animal antibacterial use and no animal to human, animal to environment, animal to animal transmission | $\Lambda_A = 0$<br>$\beta_{AH} = \beta_{AE} = \beta_{AA} = 0$ | 7.2 (1.2 – 19.5%) | 96.4 (72.3 – 99.8%) | 37.2 (4.7 – 90.6%) |
| No animal to animal, animal to human, animal to environment transmission | $\beta_{AA} = \beta_{AH} = \beta_{AE} = 0$ | 7.2 (1.2 – 19.5%) | 13.9 (1.7 – 52.7%) | 37.2 (4.7 – 90.6%) |
| No animal antibacterial use | $\Lambda_A = 0$ | 6.2 (0.8 – 19.0%) | 96.4 (72.3 – 99.8%) | 30.8 (4.1 – 81.6%) |
| No human to animal, animal to human transmission | $\beta_{HA} = \beta_{AH} = 0$ | 5.7 (0.5 – 17.4%) | 3.5 (0.3 – 21.8%) | 3.3 (0.4 – 12.7%) |
| No animal to human transmission | $\beta_{AH} = 0$ | 5.6 (0.5 – 17.2%) | 0.2 (0.0 – 2.1%) | 1.7 (0.1 – 9.5%) |
| Increasing rate of loss of resistance in animals to max | $\mu_A = 1$ | 4.6 (0.8 – 10.0%) | 71.3 (57.7 – 81.5%) | 25.5 (3.1 – 59.0%) |
| No environment antibacterial use and no environment to human, environment to environment, environment to animal transmission | $\Lambda_E = 0$<br>$\beta_{EH} = \beta_{EE} = \beta_{EA} = 0$ | 2.0 (0.1 – 6.3%) | 1.5 (0.1 – 9.0%) | 96.8 (82.3 – 99.8%) |
| No environment to animal, environment to human, environment to environment transmission | $\beta_{EA} = \beta_{EH} = \beta_{EE} = 0$ | 2.0 (0.1 – 6.3%) | 1.5 (0.1 – 9.0%) | 12.6 (0.7 – 56.3%) |
| No environment to human, environment to animal, animal to environment, human to environment transmission | $\beta_{EH} = \beta_{EA}$<br>$= \beta_{AE}$<br>$= \beta_{HE} = 0$ | 2.0 (0.1 – 6.3%) | 1.5 (0.1 – 9.0%) | 93.5 (40.7 – 99.6%) |
| No environment to human, human to environment transmission | $\beta_{EH} = \beta_{HE}$<br>$= 0$ | 1.9 (0.1 – 6.1%) | 0.5 (0.0 – 4.3%) | 39.1 (2.3 – 91.1%) |
| No environment to human transmission | $\beta_{EH} = 0$ | 1.8 (0.1 – 6.1%) | 0.1 (0.0 – 0.6%) | 0.5 (0.0 – 2.4%) |
| No environment antibacterial use | $\Lambda_E = 0$ | 1.7 (0.1 – 6.2%) | 1.4 (0.0 – 8.7%) | 96.8 (82.3 – 99.8%) |
| Increasing rate of loss of resistance in the environment to max | $\mu_E = 1$ | 1.5 (0.1 – 4.9%) | 1.2 (0.0 – 6.8%) | 82.4 (68.3 – 92.4%) |
| No animal to animal transmission | $\beta_{AA} = 0$ | 0.7 (0.0 – 5.4%) | 12.4 (1.6 – 48.6%) | 3.7 (0.2 – 25.1%) |
| No human to environment transmission | $\beta_{HE} = 0$ | 0.6 (0.0 – 3.2%) | 0.4 (0.0 – 4.2%) | 39.1 (2.3 – 91.1%) |
| No environment to animal, animal to environment transmission | $\beta_{EA} = \beta_{AE}$<br>$= 0$ | 0.6 (0.0 – 3.0%) | 1.3 (0.0 – 9.0%) | 34.2 (2.3 – 90.0%) |
| No animal to environment transmission | $\beta_{AE} = 0$ | 0.5 (0.0 – 3.0%) | 0.3 (0.0 – 4.1%) | 34.2 (2.3 – 90.0%) |
| No environment to environment transmission | $\beta_{EE} = 0$ | 0.2 (0.0 – 1.9%) | 0.1 (0.0 – 2.3%) | 11.1 (0.4 – 55.8%) |
| No human to animal transmission | $\beta_{HA} = 0$ | 0.2 (0.0 – 2.0%) | 3.5 (0.2 – 21.6%) | 1.1 (0.0 – 9.2%) |
| No environment to animal transmission | $\beta_{EA} = 0$ | 0.1 (0.0 – 0.7%) | 1.2 (0.0 – 9.0%) | 0.3 (0.0 – 3.7%) |

## References

1. O’Neill J. Tackling drug-resistant infections globally: Final report and recommendations. The review on antimicrobial resistance. London: HM Government and the Wellcome Trust; 2016.

2. WHO. Antimicrobial resistance. Global report on surveillance. World Heal Organ. 2014. doi:10.1007/s13312-014-0374-3

3. WHO. Global action plan on antimicrobial resistance. World Heal Organ. 2015.

4. Hernando-Amado S, Coque TM, Baquero F, Martínez JL. Defining and combating antibiotic resistance from One Health and Global Health perspectives. Nat Microbiol. 2019;4: 1432–1442. doi:10.1038/s41564-019-0503-9

5. Van Boeckel TP, Glennon EE, Chen D, Gilbert M, Robinson TP, Grenfell BT, et al. Reducing antimicrobial use in food animals. Science (80-). 2017;357: 1350–1352. doi:10.1126/science.aao1495

6. Van Boeckel TP, Pires J, Silvester R, Zhao C, Song J, Criscuolo NG, et al. Global trends in antimicrobial resistance in animals in low- And middle-income countries. Science (80-). 2019;365: eaaw1944. doi:10.1126/science.aaw1944

7. Holmes AH, Moore LSP, Sundsfjord A, Steinbakk M, Regmi S, Karkey A, et al. Understanding the mechanisms and drivers of antimicrobial resistance. Lancet. 2016;9: 176–187. doi:10.1016/S0140-6736(15)00473-0

8. Laxminarayan R, Matsoso P, Pant S, Brower C, Røttingen JA, Klugman K, et al. Access to effective antimicrobials: a worldwide challenge. Lancet. 2015;387: 168–175. doi:10.1016/S0140-6736(15)00474-2

9. World Health Organization. The evolving threat of antimicrobial resistance: options for action. Geneva. 2012.

10. Woolhouse M, Ward M, Van Bunnik B, Farrar J. Antimicrobial resistance in humans, livestock and the wider environment. Philos Trans R Soc B Biol Sci. 2015;370: 1–7. doi:10.1098/rstb.2014.0083

11. Lipsitch M, Samore MH. Antimicrobial use and antimicrobial resistance: A population perspective. Emerg Infect Dis. 2002;8: 347–354. doi:10.3201/eid0804.010312

12. Wernli D, Jørgensen PS, Morel CM, Carroll S, Harbarth S, Levrat N, et al. Mapping global policy discourse on antimicrobial resistance. BMJ Glob Heal. 2017;2: e000378. doi:10.1136/bmjgh-2017-000378

13. Smith E, Lichten CA, Taylor J, MacLure C, Lepetit L, Harte E, et al. Evaluation of the Action Plan against the rising threats from antimicrobial resistance. Final report. European Commission. 2016.

14. White A, Hughes JM. Critical Importance of a One Health Approach to Antimicrobial Resistance. Ecohealth. 2019;16: 404–409. doi:10.1007/s10393-019-01415-5

15. Huttner A, Harbarth S, Carlet J, Cosgrove S, Goossens H, Holmes A, et al. Antimicrobial resistance: A global view from the 2013 World Healthcare-Associated Infections Forum. Antimicrob Resist Infect Control. 2013;2. doi:10.1186/2047-2994-2-31

16. Murray CJL, Vos T, Lozano R, Naghavi M, Flaxman AD, Michaud C, et al. Disability-adjusted life years (DALYs) for 291 diseases and injuries in 21 regions, 1990-2010: a systematic analysis for the Global Burden of Disease Study 2010. Lancet. 2012;380: 2197–2223. doi:10.1016/S0140-6736(12)61689-4

17. Allegranzi B, Nejad SB, Combescure C, Graafmans W, Attar H, Donaldson L, et al. Burden of endemic health-care-associated infection in developing countries: Systematic review and meta-analysis. Lancet. 2011;377: 228–241. doi:10.1016/S0140-6736(10)61458-4

18. Karanika S, Karantanos T, Arvanitis M, Grigoras C, Mylonakis E. Fecal Colonization with Extended-spectrum Beta-lactamase-Producing Enterobacteriaceae and Risk Factors among Healthy Individuals: A Systematic Review and Metaanalysis. Clin Infect Dis. 2016;63: 310–318. doi:10.1093/cid/ciw283

19. Niewiadomska AM, Jayabalasingham B, Seidman JC, Willem L, Grenfell B, Spiro D, et al. Population-level mathematical modeling of antimicrobial resistance: A systematic review. BMC Med. 2019;17: 1–20. doi:10.1186/s12916-019-1314-9

20. Sumpradit N, Wongkongkathep S, Poonpolsup S, Janejai N, Paveenkittiporn W, Boonyarit P, et al. New chapter in tackling antimicrobial resistance in Thailand. BMJ. 2017;358. doi:10.1136/bmj.j2423

21. Phumart P, Phodha T, Thamlikitkul V, Riewpaiboon A, Prakongsai P, Limwattanon S. Health and economic impacts of antimicrobial resistance in Thailand. J Health Serv Res Policy. 2012;6: 352–360.

22. Pongpech P, Naenna P, Taipobsakul Y, Tribuddharat C, Srifuengfung S. Prevalence of extended-spectrum beta-lactamase and class 1 integron integrase gene intl1 in Escherichia coli from thai patients and healthy adults. Southeast Asian J Trop Med Public Health. 2008;39: 425–433.

23. Luvsansharav UO, Hirai I, Nakata A, Imura K, Yamauchi K, Niki M, et al. Prevalence of and risk factors associated with faecal carriage of CTX-M β-lactamase-producing enterobacteriaceae in rural Thai communities. J Antimicrob Chemother. 2012;67: 1769–1774. doi:10.1093/jac/dks118

24. Boonyasiri A, Tangkoskul T, Seenama C, Saiyarin J, Tiengrim S, Thamlikitkul V. Prevalence of antibiotic resistant bacteria in healthy adults, foods, food animals, and the environment in selected areas in Thailand. Pathog Glob Health. 2014;108: 235–245. doi:10.1179/2047773214Y.0000000148

25. Ministry of Public Health, Ministry of Agriculture and Cooperatives. National Strategic Plan on Antimicrobial Resistance 2017-2021, Thailand. 2016. doi:10.2471/BLT.16.179648

26. van Bunnik BAD, Woolhouse MEJ. Modelling the impact of curtailing antibiotic usage in food animals on antibiotic resistance in humans. R Soc Open Sci. 2017;4: 161067. doi:10.1098/rsos.161067

27. Pal C, Bengtsson-Palme J, Kristiansson E, Larsson DGJ. The structure and diversity of human, animal and environmental resistomes. Microbiome. 2016;4. doi:10.1186/s40168-016-0199-5

28. Sasaki T, Hirai I, Niki M, Nakamura T, Komalamisra C, Maipanich W, et al. High prevalence of CTX-M β-lactamase-producing enterobacteriaceae in stool specimens obtained from healthy individuals in Thailand. J Antimicrob Chemother. 2010;65: 666–668. doi:10.1093/jac/dkq008

29. Luvsansharav UO, Hirai I, Niki M, Sasaki T, Makimoto K, Komalamisra C, et al. Analysis of risk factors for a high prevalence of extended-spectrum β-lactamase-producing Enterobacteriaceae in asymptomatic individuals in rural Thailand. J Med Microbiol. 2011;60: 619–624. doi:10.1099/jmm.0.026955-0

30. Thamlikitkul V, Tiengrim S, Thamthaweechok N, Buranapakdee P, Chiemchaisri W. Contamination by Antibiotic-Resistant Bacteria in Selected Environments in Thailand. Int J Environ Res Public Health. 2019;16: 1–11.

31. Kristiansson E, Fick J, Janzon A, Grabic R, Rutgersson C, Weijdegård B, et al. Pyrosequencing of antibiotic-contaminated river sediments reveals high levels of resistance and gene transfer elements. PLoS One. 2011;6: e17038. doi:10.1371/journal.pone.0017038

32. Knight GM, Costelloe C, Deeny SR, Moore LSP, Hopkins S, Johnson AP, et al. Quantifying where human acquisition of antibiotic resistance occurs: A mathematical modelling study. BMC Med. 2018;16: 1–11. doi:10.1186/s12916-018-1121-8

33. Tacconelli E, Carrara E, Savoldi A, Harbarth S, Mendelson M, Monnet DL, et al. Discovery, research, and development of new antibiotics: the WHO priority list of antibiotic-resistant bacteria and tuberculosis. Lancet Infect Dis. 2018;18: 318–327. doi:10.1016/S1473-3099(17)30753-3

34. Coyne L, Arief R, Benigno C, Giang VN, Huong LQ, Jeamsripong S, et al. Characterizing antimicrobial use in the livestock sector in three south east asian countries (Indonesia, Thailand, and Vietnam). Antibiotics. 2019;8: 33. doi:10.3390/antibiotics8010033

35. Khamsarn S, Nampoonsak Y, Busamaro S, Tangkoskul T, Seenama C, Rattanaumpawan P, et al. Epidemiology of antibiotic use and antimicrobial resistance in selected communities in Thailand. J Med Assoc Thail. 2016;99: 1–6.

36. Ashley EA, Shetty N, Patel J, Van Doorn R, Limmathurotsakul D, Feasey NA, et al. Harnessing alternative sources of antimicrobial resistance data to support surveillance in low-resource settings. J Antimicrob Chemother. 2019;74: 541–546. doi:10.1093/jac/dky487

